# SAA positivity rate amongst dual LRRK2-GBA1, GBA1 and LRRK2 carriers with Parkinson’s disease

**DOI:** 10.64898/2026.05.26.26354106

**Authors:** Penina Ponger, Anuprita R Nair, Noah S Johnson, Chelsea Caspell-Garcia, David-Erick Lafontant, Roy N Alcalay, The Parkinson’s Progression Markers Initiative

## Abstract

We investigated whether people with Parkinson’s disease who are dual GBA1+LRRK2 carriers have a milder, LRRK2-like phenotype as previously reported. This was accomplished by comparing clinical features and alpha-synuclein seed amplification assay (SAA) positivity rates between dual GBA1+LRRK2-PD(n=13), GBA1-PD(n=169) and LRRK2-PD(n=175) carriers in a cross-sectional retrospective study of Parkinson’s Progression Markers Initiative (PPMI) data. Our results show that GBA1+LRRK2-PD SAA positivity rate(83%) is closer to GBA1-PD rate(87%) rather than LRRK2-PD rate(62%,p-value>0.05). GBA1+LRRK2-PD have both non-motor and motor phenotypic similarity to GBA1-PD,(p-value>0.05). This small PPMI cohort indicates that dual GBA1+LRRK2-PD carriers’ SAA positivity and phenotype are aligned with GBA1-PD.

## Introduction

Parkinson’s disease (PD) is the second most common neurodegenerative disease, affecting approximately 2% of people over 65 worldwide[1] with variability in onset, progression rate and prodromal, non-motor, and motor findings.[2,3] Attaining clear diagnostic criteria and providing tailored therapy remain difficult in light of such phenotypic variability. Further validation of biomarkers, including alpha-synuclein seed amplification assays (SAA) and profiling genetic risk factors aid in bridging these gaps.[4-8]

Genetic variants in *GBA1* and *LRRK2* represent the most common PD risk factors, each gene presenting with distinct clinical and biological features. *GBA1*-associated PD (GBA1-PD) is characterized by increased non-motor involvement, including hyposmia, REM sleep behavior disorder (RBD), mood disturbances, and increased risk of dementia.*LRRK2*-associated PD (LRRK2-PD) has slower progression with fewer non-motor features. Importantly, dual GBA1+LRRK2 carriers have been reported to present with a milder clinical course than GBA1-PD alone, suggesting a possible protective interaction between these two genetic risk factors. The biological basis for such an assumption remains unclear.[4-5,9-10]

SAA has emerged as a highly sensitive and specific biomarker for alpha-synuclein pathology in PD and its prodromal states.[11-13] Prior studies have shown that nearly all GBA1-PD patients have positive SAA rate, compared to only two-thirds of LRRK2-PD. The prevalence and clinical correlates of SAA positivity rate in dual GBA1+LRRK2 carriers, however, remain unknown. Understanding whether SAA levels in this subgroup align more closely with GBA1-PD, LRRK2-PD, or demonstrate an intermediate pattern is critical for elucidating the interaction between these genes and their contribution to PD pathogenesis.

In this study, leveraging data from the Parkinson’s Progression Markers Initiative (PPMI), we investigate SAA status among dual GBA1+LRRK2 carriers and provide new insights into how genetic risk factors interact to shape PD pathophysiology.[14]

## Methods

### Overview of Study

PPMI is a longitudinal study sponsored by The Michael J. Fox Foundation for Parkinson’s Research (MJFF), recruiting participants since 2010. SAA samples have been collected since 2022. This study was designed as a cross-sectional retrospective study. All study activities were approved by the Western-Copernicus Group institutional review board and informed consent was obtained from participants before participation.

### Participants

PPMI data (www.ppmi-info.org/access-data-specimens/download-data, RRID:SCR 006431) queried on February 3, 2025, was extracted for all individuals with PD who were either *LRRK2-PD, GBA1-PD* or dual GBA1+LRRK2-PD carriers with SAA status. Participants were genotyped using either Next Generation Sequencing for LRRK2 and GBA1 variants and via NeuroX-derived genotyping or the Immunochip Array for GBA1 variants. Homozygous GBA1 carriers were excluded. Carriers of two GBA1 variants were included.[15,16]

### Statistical Analysis

Statistical analyses were performed using SAS v9.4 (SAS Institute Inc.,Cary, NC;sas.com; RRID:SCR_008567). All tests were 2-sided, and *p-value*<0.05 was considered significant.

The SAA positivity rates amongst the three genetic PD subgroups were assessed. A cluster bar graph was plotted to represent percentages of positive, negative, Inconclusive and MSA-like SAA categories across the genetic subgroups. Statistical comparison of SAA positivity between the subgroups was performed using Chi-square or Fisher’s exact tests as appropriate with reported p-values from these tests.

Descriptive statistics at baseline were presented for the PD participants across the three genetic subgroups, including frequencies (percentages) for categorical measures and medians and interquartile ranges (IQRs) for continuous measures. Pairwise comparisons between the three subgroups were performed using Wilcoxon rank-sum test for continuous variables, and Chi-square or Fisher’s exact test for categorical variables with reported p-values from these tests.

## Results

### Demographic and clinical characteristics

A total of 357 participants with PD participated in this study. Mean age was 62.9 years [9.5 SD],174 (48.7%) were women and the overall disease duration for the cohort was 2.5 years [2.2SD]. Genetic subgroups included 13 GBA1+LRRK2-PD, 169 GBA1-PD and 175 LRRK2-PD.(Table 1) GBA1+LRRK2-PD participants were younger at enrolment (58 years compared to 62.9 in GBA1-PD and 65.2 in LRKK2-PD), age of onset (53.8 years compared to 59.5 for GBA1-PD and 60.1 for LRRK2-PD) and diagnosis of disease (54.7 years compared to 60.8 for GBA1-PD and 61.6 for LRRK2-PD). GBA1+LRRK2-PD included 85% females as opposed to nearly equal representation of sex in the two other groups (p<0.01). Dual carriers had longer disease duration (4.0 years) compared to GBA1-PD (1.2 years,p<0.01) and LRRK2-PD (2.3 years, without statistical significance). See figures 1A-C and Supplement 1 for genetic variants.

**Figure 1A:**
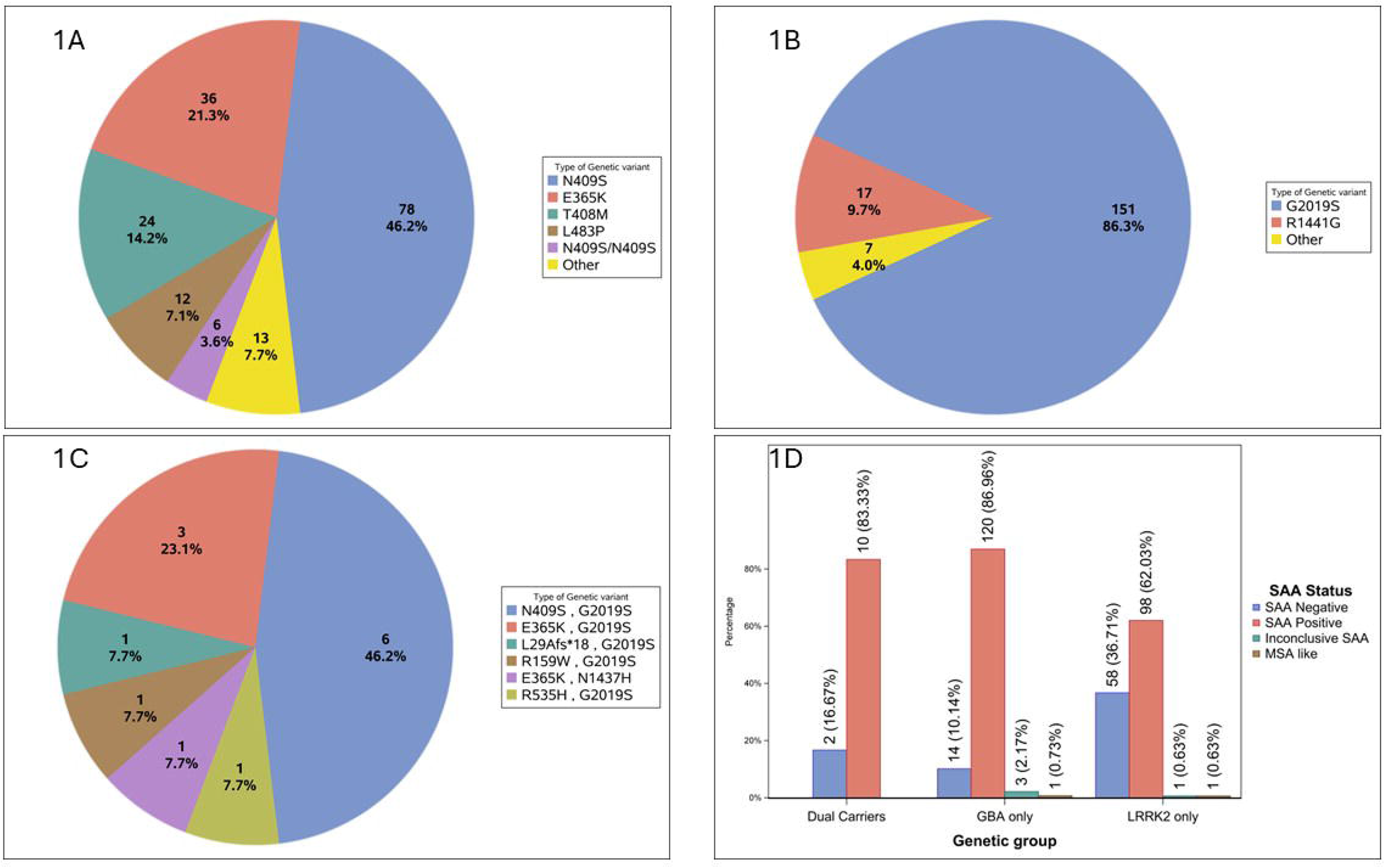
Distribution of GBA1-PD variants; ‘Other’ category includes L29Afs*18(N=3), IVS2+1G>A(N=2), N409S/E365K(N=2), F216Y(N=1), IVS2+1G>A/T408M(N=1), L483P/E365K(N=1), N409S / c.762-2A>G(N=1), R159W/T408M(N=1), and R502C(N=1) GBA1 variants. Figure 1B) Distribution of LRRK2-PD variants; ‘Other’ category includes G2019S/G2019S(N=5), I2020T(N=1), R1441C(N=1) LRRK2 variants. Figure 1C: Distribution of GBA1+ LRRK2-PD variants. Figure 1D: SAA positivity across genetic groups in the PD cohort.

#### SAA positivity rate

Dual carriers had a SAA positivity rate of 83%, which was markedly closer to GBA1-PD rate of 86% compared to the LRRK2-PD rate of 62%. This finding was without statistical significance. SAA positivity rate was significantly (p<0.01) higher amongst GBA1-PD carriers compared to LRRK2-PD carriers.(Figure 1D)

#### Motor features

The median MDS-UPDRS III(ON) scores ranged from 16 for LRRK2-PD, 18 in the GBA1+LRRK2-PD group and 21 in the GBA1-PD group with statistical significance between the single gene carriers (p<0.01). GBA1-PD and dual carriers’ group had higher MDS-UPDRS III (OFF) scores (22) compared to LRRK2-PD group (18) but data availability for OFF state was partial in all groups.

#### Non motor features

The groups had similar MOCA measures (27). UPSIT percentile scores were lower amongst GBA1-PD (4.8) and GBA1+LRRK2-PD (5.0) groups with statistical significance between the LRRK2-PD (11.0) and GBA1-PD groups (p<0.01). SCOPA-AUT scores were higher in GBA1+LRRK2-PD (14) compared to the two other groups (10,p>0.05).

#### Biomarkers

Urine total di-18:1-BMP levels were higher in both the LRRK2-PD (12.1 IQR) and GBA1+LRRK2-PD (16.4 IQR) group with statistical significance between the LRRK2-PD and GBA1-PD groups (4.7 IQR,p<0.01).

## Discussion

Our cross-sectional study shows that SAA positivity rate amongst GBA1+LRRK2-PD approximates that observed in GBA1-PD rather than LRRK2-PD, although statistical significance was not reached. Consistently, dual carriers exhibited motor, non-motor, and biomarker profiles that overlapped with those of GBA1-PD, with several associations reaching partial statistical significance. These findings indicate that dual GBA1+LRRK2-PD is characterized by a predominantly GBA1-like phenotype.

This unexpected result contrasts with prevailing models, which have largely portrayed dual GBA1+LRRK2-PD as phenotypically milder and more closely aligned with LRRK2-PD, suggesting that LRRK2 variants exert a dominant or protective modifying effect on GBA1-associated disease, attenuating motor and non-motor manifestations.[12-13]

In a retrospective cohort, consisting of 236 PD patients including 66 LRRK2-PD, 77 GBA-PD and 12 GBA1+LRRK2-PD patients, Yahalom and colleagues reported an increased PD risk among GBA1+LRRK2-PD with estimated odds ratio (OR) risk for PD in GBA1+LRRK2-PD being 28 ( 95% CI,10.9–72.0;p < 0.0001) compared to 2.7–3.5 OR in GBA-PD and 8.6 OR in LRRK2-PD. The group proposed an additive genetic effect, while observing trends toward younger age at onset and less severe non-motor features relative to GBA1-PD, leading to the hypothesis that LRRK2 variants may partially offset the deleterious impact of GBA1 variants.[17]

Omer and colleagues, in their prospective study analyzing phenotypic features of 1090 patients with idiopathic PD, 155 LRRK2-PD, 211 GBA1-PD, and 27 GBA1+LRRK2-PD patients, reported lower UPDRS scores and preserved olfaction in LRRK2-PD and GBA1+LRRK2-PD compared with GBA1-PD, further supporting the notion of LRRK2-driven phenotypic dominance in GBA1+LRRK2-PD.[18]

Longitudinal data from Ortega and colleagues provided only partial support for this paradigm. In their study of 1193 participants, (128 *GBA1-*PD, 155 L*RRK2-*PD and 21 GBA1+LRRK2-PD), GBA1+LRRK2-PD demonstrated slower cognitive decline than GBA1-PD, thus resembling LRRK2-PD patients. On the other hand, motor progression in GBA1+LRRK2-PD did not significantly differ from that observed in GBA1-PD. Accordingly, the study did not demonstrate a significant interaction between *LRRK2* G2019S and *GBA1* variants effecting patients’ motor progression. This larger study indicates a possible paradigm shift regarding GBA1+LRRK2-PD phenotype.[19]

Our report is the first, to the best of our knowledge, that describes both clinical and biological biomarker features amongst PD patients according to their genetic status, providing preliminary evidence that LRRK2 and GBA1 variant interaction differs from prior reports. We demonstrate that amongst GBA1+LRRK2-PD motor, non motor and SAA positivity behave similarly to patterns seen in GBA1-PD.

Our study is limited by a small sample size and cross-sectional design. Prospective longitudinal data will firstly, enable analysis of evolving variables from initially non manifesting carriers and their clinical conversion patterns and secondly, delineate distinct SAA kinetic profiles reflecting PD progression rate, including patterns of cognitive decline. Larger cohorts will enable multi-analysis of clinical features.

In addition, heterozygote carriers of all known GBA1 variants were included in this study whereas all homozygous carriers were excluded. All selected genetic variants are known to be contributing PD risk factors yet our current knowledge of the significance of low risk GBA1 variants and homozygote carrier status remains limited. These genetic variables may have implications on PD patient’s phenotypes, resulting in selection bias in this study.[20-21]

Furthermore, the cohort included carriers of two GBA1 variants, which may be compound heterozygote carriers if variants are in trans. This could potentially contribute to increased SAA positivity in this group due to dosage effect.[20-21] Our GBA1+LRRK2-PD carrier group also had longer disease duration which could also affect the SAA positivity rate. Lastly, the dual carrier group in this study was predominantly female, which may affect both non motor and motor PD features.[22]

Notwithstanding these limitations, our findings highlight the need for large, longitudinal, biomarker-informed studies to refine models of GBA1–LRRK2 interaction. Such efforts will be essential to accurately define disease mechanisms, optimize genetic stratification, and inform precision therapeutic approaches in PD.

## Supporting information

Genetic variables table

PPMI authors list

## Data Availability

www.ppmi-info.org/access-data-specimens/download-data

https://zenodo.org/records/18189221?preview=1&token=eyJhbGciOiJIUzUxMiJ9.eyJpZCI6IjNjZWFkOTdjLTk1Y2YtNDMwMC04NzNlLTk4Yzg5MTI5NjQ0MCIsImRhdGEiOnt9LCJyYW5kb20iOiJlZDhmMGFlYzNlOTcwOTY3N2JjOTdmY2YxNjcyYTFkYSJ9.5KCBNt1FLoc0PQEDADOFwSVd6oNR6u_L3hKJ537Pz5EdCe8lw4bMxBJHkEYxrnGBceCTVH3GYOj6CjubvQVt4g

## Acknowledgements

The authors express their sincere gratitude to all participants with PD for their invaluable participation in the PPMI study.

## Authors’ Roles

(1) Research project: A. Conception, B. Organization, C. Clinical Examination, D. Execution and Data Analysis; (2) Statistical Analysis: A. Design, B. Execution, C. Review and Critique; (3) Manuscript Preparation: A. Writing of the First Draft, B. Review and Critique.

P.P. 1B 1C 1D 2A 2C 3A 3B

A.N. 2B 3A 3B

N.J. 2B 3B

C.C.G. 2A 2C 3B

D.E.L. 2A 2C 3B

R.A. 1A, 2C, 3B

## Table and Figures Legend

Table 1: Demographic and Clinical Features according to Genetic Status

**Supplement 1:** Distribution of genetic variants for cohort

**Supplement 2:** PPMI Author List for Publications

