## Supplementary material for "SAA positivity rate amongst dual LRRK2-GBA1, GBA1 and LRRK2 carriers with Parkinson’s disease": Genetic variables table

- Distribution of participants across the PD cohort (N=357)

| **Group** | **N (%)** |
| --- | --- |
| **Dual Carriers** | 13 (3.7%) |
| **GBA1 only** | 169 (47.3%) |
| **LRRK2 only** | 175 (49.0%) |

- Distribution of genetic variants among GBA1 only group (N=169)

| **GBA1 variant** | **N (%)** |
| --- | --- |
| **E365K** | 36 (21.3%) |
| **F216Y** | 1 (0.6%) |
| **IVS2+1G>A** | 2 (1.2%) |
| **IVS2+1G>A/T408M** | 1 (0.6%) |
| **L29Afs*18** | 3 (1.8%) |
| **L483P** | 12 (7.1%) |
| **L483P/E365K** | 1 (0.6%) |
| **N409S** | 78 (46.1%) |
| **N409S / c.762-2A>G** | 1 (0.6%) |
| **N409S/E365K** | 2 (1.2%) |
| **N409S/N409S** | 6 (3.5%) |
| **R159W/T408M** | 1 (0.6%) |
| **R502C** | 1 (0.6%) |
| **T408M** | 24 (14.2%) |

- Distribution of genetic variants among LRRK2 only group (N=175)

| **LRRK2 variant** | **N (%)** |
| --- | --- |
| **G2019S** | 151 (86.3%) |
| **G2019S/G2019S** | 5 (2.8%) |
| **I2020T** | 1 (0.6%) |
| **R1441C** | 1 (0.6%) |
| **R1441G** | 35 (9.7%) |

- Distribution of genetic variants among Dual Carriers group (N=13)

| **Dual Carriers** | |
| --- | --- |
| **Genetic variants** | **N (%)** |
| **E365K + G2019S** | 3 (23.1%) |
| **E365K + N1437H** | 1 (7.7%) |
| **L29Afs*18 + G2019S** | 1 (7.7%) |
| **N409S + G2019S** | 6 (46.1%) |
| **R159W + G2019S** | 1 (7.7%) |
| **R535H + G2019S** | 1 (7.7%) |
