## Supplementary figures and images for "SAA positivity rate amongst dual LRRK2-GBA1, GBA1 and LRRK2 carriers with Parkinson’s disease"

### PPMI authors list

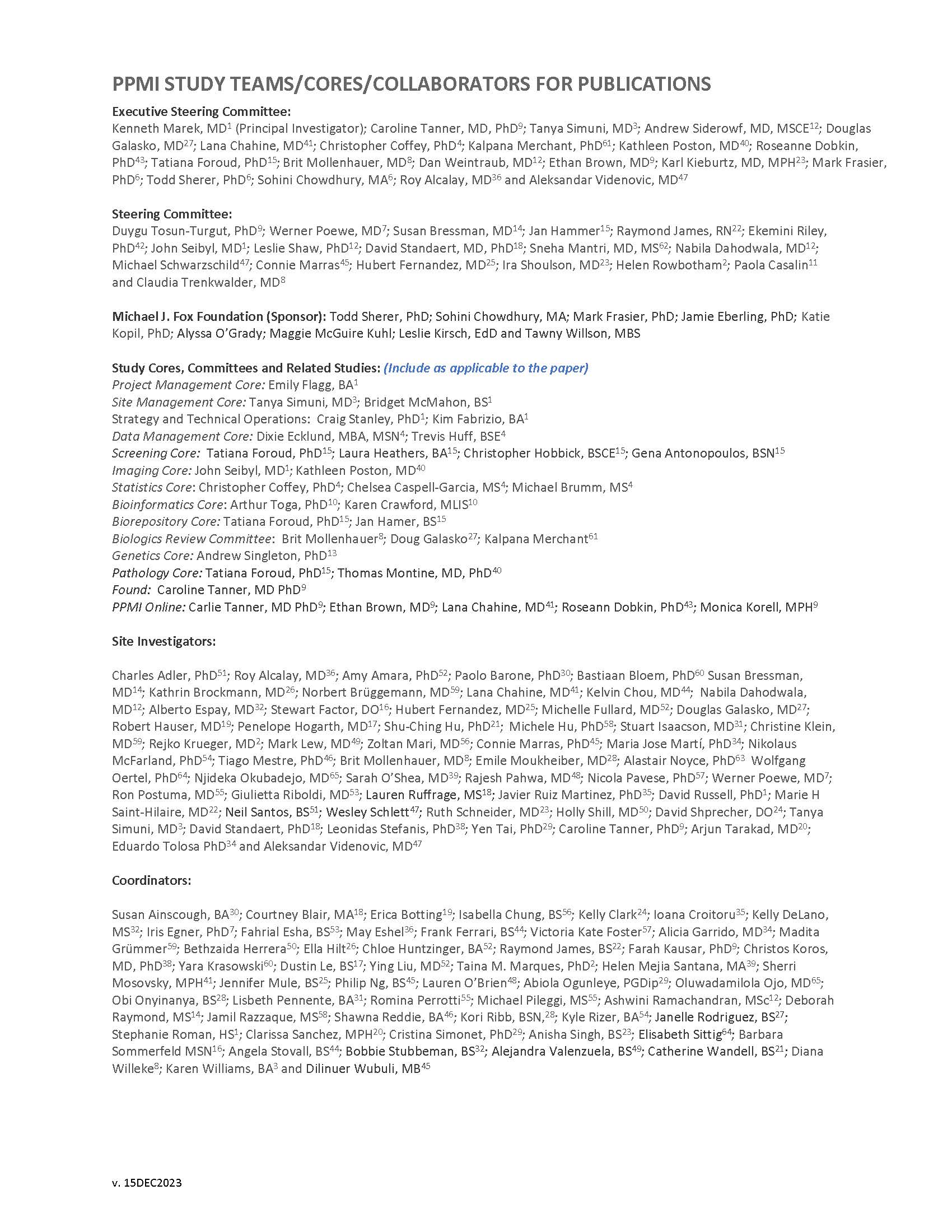
